# Integrating Local Vocabulary into OMOP CDM: A Step-by-Step Tutorial

**DOI:** 10.1101/2025.05.07.25327200

**Authors:** Yiju Park, Jinwoo Yoon, Aleh Zhuk, Anna Ostropolets, Seng Chan You

## Abstract

**Objectives:** This tutorial provides healthcare informatics researchers with a practical guide for integrating local vocabularies into the Observational Health Data Sciences and Informatics (OHDSI) Standardized Vocabularies, through the example of Korean Electronic Data Interchange (EDI) codes.

**Methods:** We present a step-by-step guide for integrating local vocabulary. The methodology includes four sequential stages: (1) comprehensive and continuous collection system for longitudinal EDI codes from November 2000 to May 2024, (2) mapping to OHDSI Standardized Vocabularies, (3) validation and integration through the OHDSI Community Contribution Pipeline, and (4) publication into the Athena, the official OHDSI vocabulary site.

**Results:** By following this tutorial, researchers can achieve integration of local vocabulary into the OHDSI Vocabularies. Of the total 620,642 EDI codes collected, we successfully integrated 437,807 codes (70.5%) into the OHDSI Vocabularies, with 376,111 (85.9%) of these integrated codes mapped to standard concepts. The integrated vocabulary passed all Community Contribution Pipeline requirements and was published in September 2024 on the official OHDSI vocabulary website.

**Conclusion:** This tutorial provides healthcare informatics researchers with practical guidance for vocabulary standardization, enabling local healthcare data integration into the OMOP CDM. The approach can be adapted for various local vocabularies, fostering international healthcare data interoperability.

## INTRODUCTION

The standardization of healthcare data across different systems is essential for enabling large-scale observational research using real-world data[1]. The Observational Medical Outcomes Partnership Common Data Model (OMOP CDM), maintained by the Observational Health Data Sciences and Informatics (OHDSI), has emerged as a widely adopted framework for standardizing healthcare data structure and harmonizing its content[2]. As of 2024, over 540 data sources from 54 countries have been standardized to OMOP CDM[3]. A critical challenge in CDM adoption is incorporating local vocabularies used in different healthcare systems into the OHDSI Standardized Vocabularies, which is a mandatory reference standard in OMOP CDM. Vocabularies represent a collection of source terminologies, ontologies and vocabularies and crosswalks to reference standard terminologies. Integration of the local vocabularies into the OHDSI Vocabularies provides benefits for standardized and scalable research but involves multiple complex steps that require both technical expertise and domain knowledge.

The OHDSI community recently introduced the Community Contribution Pipeline[4], establishing standardized guidelines for vocabulary integration. These guidelines primarily emphasize validation aspects while providing less detailed instruction on the comprehensive process from local vocabulary collection through mapping methodologies leaving such aspects to contributors. This presents additional challenges for newcomers seeking end-to-end guidance on vocabulary integration.

Our research group previously integrated Korean EDI codes into the OMOP CDM in 2020, using available vocabulary at a single time point (October 2019)[5]. While this initial effort was valuable, it achieved limited mapping coverage and lacked a comprehensive approach for handling the longitudinal nature of vocabulary systems.

This tutorial aims to provide a practical guide for integrating local vocabulary into the OHDSI Vocabularies using a semi-automated approach, with Korean Electronic Data Interchange (EDI) codes as a case example. We present a comprehensive step-by-step process covering the entire workflow from initial data collection, through data processing, mapping to international standard terminologies, evaluation and validation of terms to be loaded, to the final publication of terminology on vocabulary sites. The methodology presented here is designed to be accessible to beginners while providing sufficient detail for adaptation to other local vocabularies, thereby enhancing healthcare data interoperability across diverse healthcare systems.

## DESCRIPTION

We developed a semi-automated pipeline consisting of four main components (Figure 1). Our semi-automated approach automates data collection, processing, validation, and publication steps, while the integration with standard concepts requires manual expert review to ensure accuracy and semantic consistency. This combination of automation and expert judgment optimizes both efficiency and quality in vocabulary integration. Full details of all systems are given in the supplementary information.

**Figure 1.**
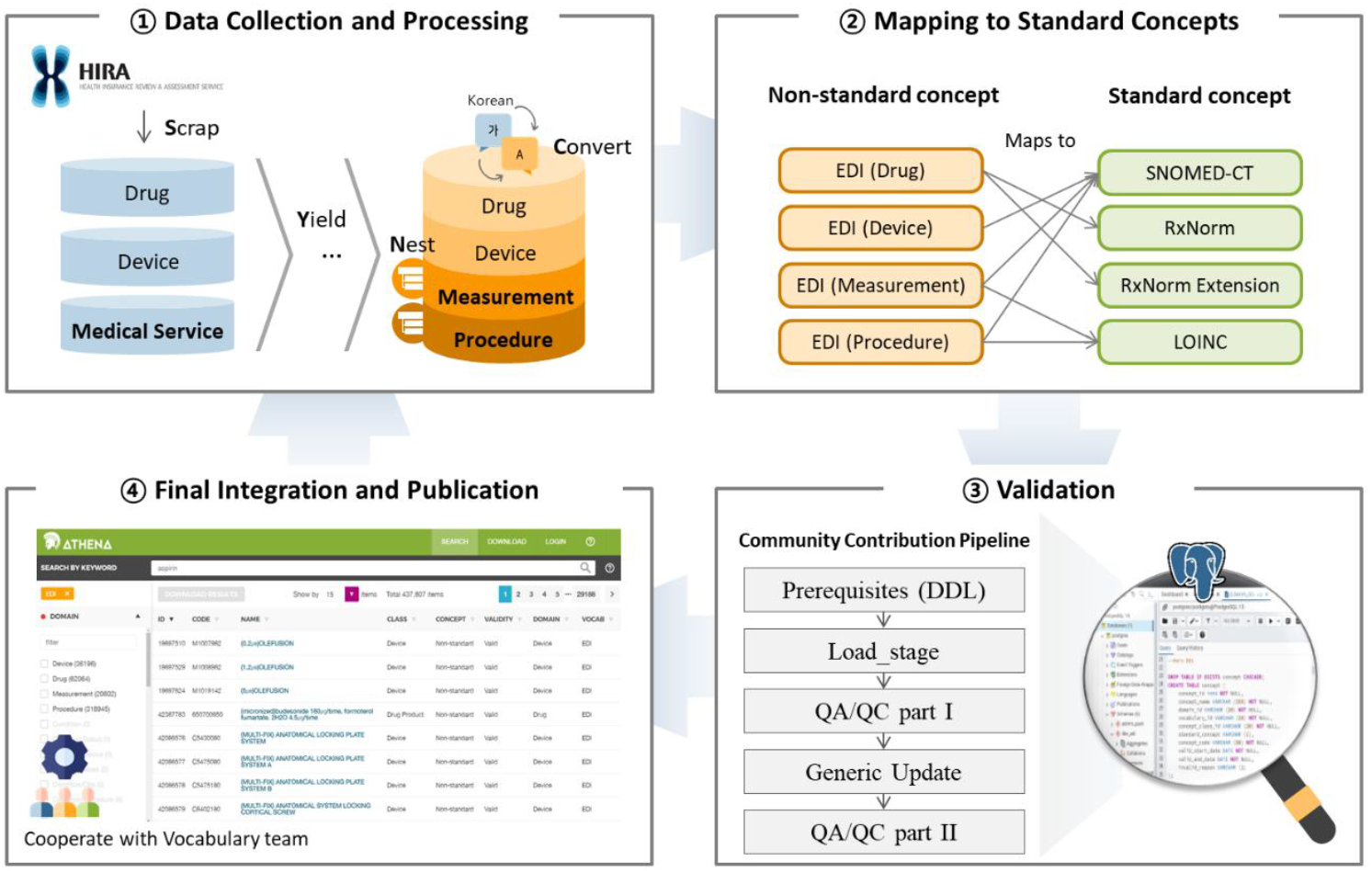
The overall process of EDI vocabulary integration into OMOP CDM

### Stage 1: Data Collection

#### Data Source

For this study, we collected EDI codes across all domains (Drug, Device, and Medical Service) from November 2000 to May 2024. In Korea, the EDI vocabulary system is the standard coding system for healthcare claims and electronic health records, provided by the Health Insurance Review & Assessment Service (HIRA).

While widely used in Korea, EDI vocabulary presents several challenges for standardization efforts. First, HIRA releases updates to the EDI codes monthly for each domain separately, requiring frequent downloads and continuous maintenance. Additionally, the system lacks concept permanence, meaning expired codes can be reassigned to different concepts over time (re-used). The system also suffers from semantic inconsistencies, where the same codes may have different meanings depending on the context or the time period. These characteristics make the EDI vocabulary particularly challenging to harmonize with international standard terminologies.

#### Data Processing

Local vocabulary often requires comprehensive and continuous system approaches due to frequent updates, semantic inconsistencies, and longitudinal characteristics. Considering EDI’s longitudinal characteristics and code duplications, we implemented a comprehensive and continuous collection system, utilizing ‘SYNC’[6] (a modified version of previous ‘EdiToOmop’ package[5, 7]) as a tool to track code updates and deprecations over the 23-year period. Figure 2 shows examples of how this system tracks code updates and deprecations. Our local vocabulary collection system performs four sequential steps (Figure 3):

**Figure 2.**
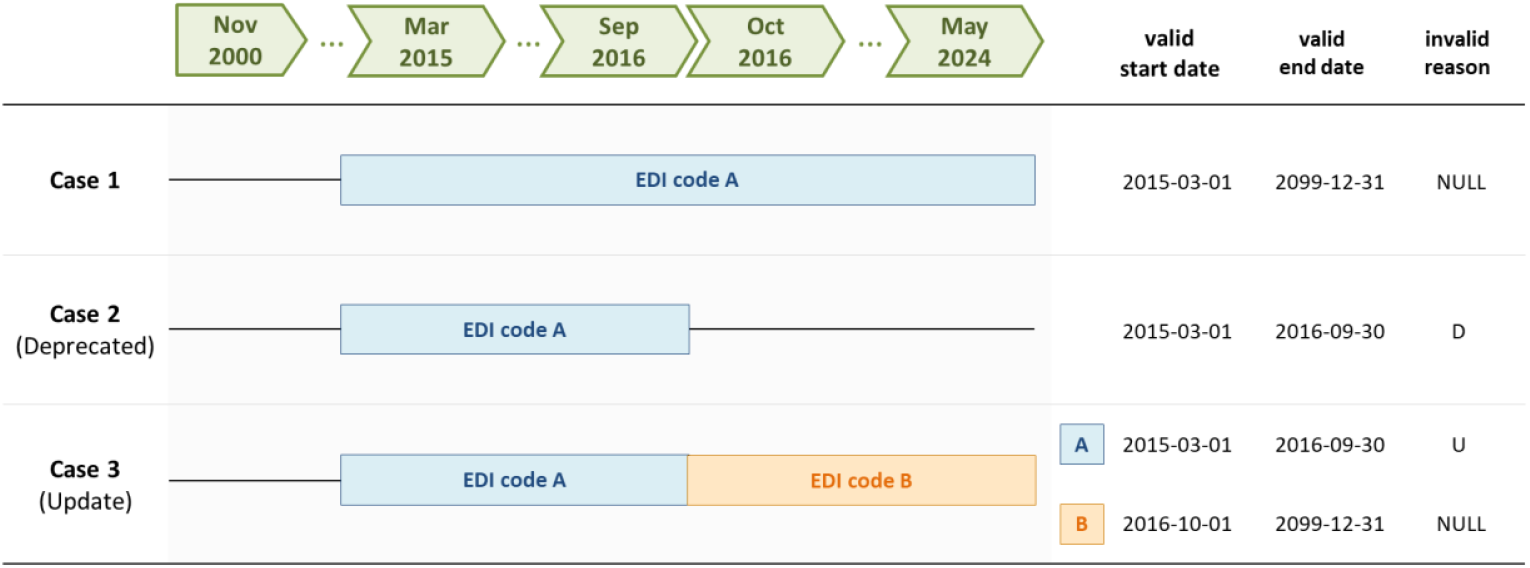
Examples of EDI codes type

**Figure 3.**
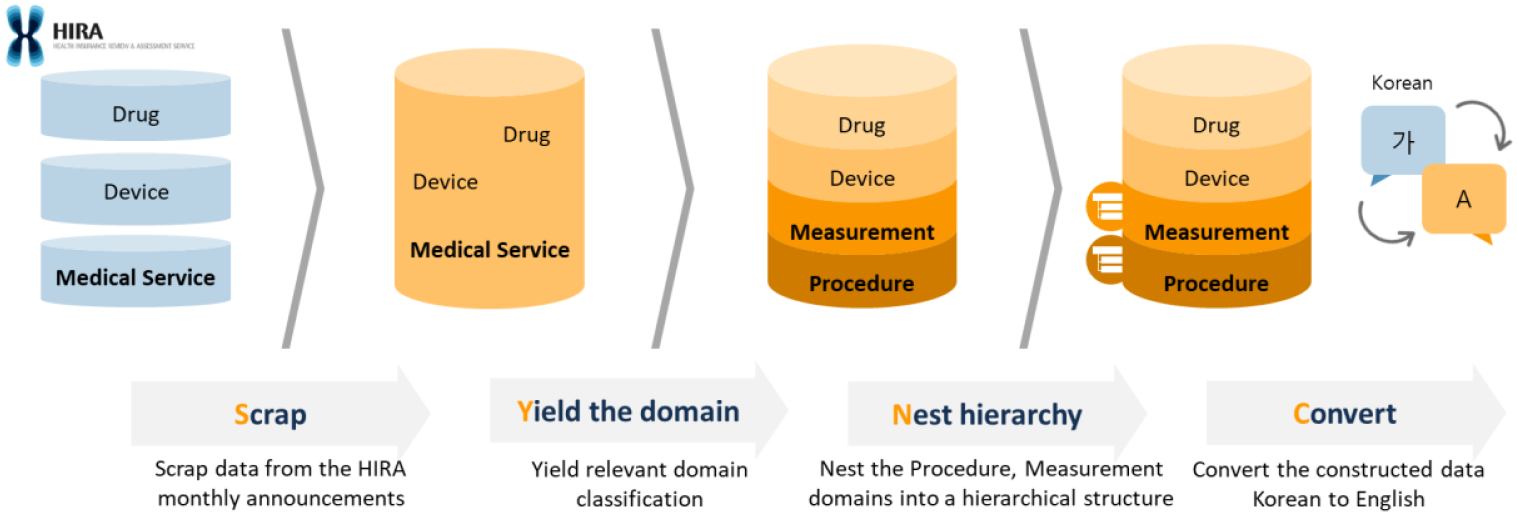
The process of the semi-automated ‘SYNC’ package

1. Scraping: Systematic extractsion of data from HIRA’s monthly announcements (Drug[8], Device, and Medical Service[9])
2. Yielding: Datasets were classified into four domains corresponding to the OMOP CDM domains - Device, Drug, Procedure, and Measurement. While the original EDI
3. data only distinguished three domains (Device, Drug, and Medical Service), we further subdivided Medical Service codes into Procedure and Measurement domains based on their characteristics Nesting: Hierarchical structuring of domains (e.g., assigning Procedure/Proc Hierarchy concept class based on 5-digit numbers of EDI codes for Procedure domain classification)
4. Converting: Automated English translation using a commercial machine translation API[10]

The system is publicly available on GitHub[6] for transparency and reproducibility.

### Stage 2: Mapping to standard concepts

Following the domain classification through our collection system, we mapped the EDI codes to standard OMOP concepts using the “Maps to” relationship. This mapping stage is the only step in our pipeline that requires significant manual effort, as it demands both clinical domain expertise and understanding of terminology standards. For each domain, we identified appropriate standard vocabularies: RxNorm and RxNorm Extension were used for Drug domain; SNOMED CT was selected for Device domain mapping; and both SNOMED CT and LOINC were utilized for Procedure and Measurement domains to ensure comprehensive coverage of clinical procedures and laboratory tests[11]. We prioritized the Drug and Device domains based on their importance in observational studies. To ensure accuracy and consistency, the experts followed OHDSI Korea Mapping guidelines[12].

### Stage 3: Validation

The Community Contribution Pipeline is the OHDSI’s standardized process for integrating new vocabularies into the OHDSI Vocabularies, ensuring quality and consistency through automated validation steps. The pipeline[4] performs five sequential stages. Table 1 provides detailed descriptions and function lists for each stage of the Community Contribution Pipeline.

**Table 1.** List of Community Contribution Pipeline.

| Description | Functions |
| --- | --- |
| **(1) Prerequisites** | - |
| • Create extension: pg_trgm, tablefunc, plpython3u<br>• Create schemas: sources, devv5, dev_edi, qa_tests, vocabulary_pack, admin_pack<br>• Create functions |  |
| **(2) Load Stage** |  |
| • Execute load_stage.sql<br>• Load Stage commonly contains the scripts to stage the vocabulary into the common format and references two types of PostgreSQL procedures that will need to create locally | set_latest_update,<br>process_manual_synonyms,<br>process_manual_relationships,<br>add_fresh_maps_to,<br>get_actual_concept_info,<br>deprecate_wrong_maps_to,<br>delete_ambiguous_maps_to,<br>check_stage_tables |
| **(3) QA/QC Part I** |  |
| • Execute create_qa_tests.sql<br>• QA/QC Part I performs conformance checks and should return no errors, including no duplicates, no exceed field length limits, etc |  |
| **(4) Generic Update** | generic_update |
| • Execute generic_update.sql<br>• Generic Update function integrates the content of the stage tables into basic tables (CONCEPT, CONCEPT_RELATIONSHIP, etc) and assigns concept ids |  |
| **(5) QA/QC Part II** | get_summary, get_checks |
| • Execute create_qa_tests.sql<br>• Execute manual_checks_after_generic_update.sql<br>• QA/QC Part II is done semi-automatically, including automatically check the compliance of resulting basic tables to the OMOP rules, generate high-level statistics between the development and devv5 schema for CONCEPT and CONCEPT_RELATIONSHIP table |  |

All function names have been standardized from camelCase to snake\_case format for consistency.

1. Prerequisites (Environment Setup): Configuration of PostgreSQL database with three schemas – source, reference, and development – for systematic vocabulary management and vocabulary processing
2. Load Stage (Vocabulary Loading): Integration of EDI codes collected through our comprehensive collection system and their mapped with standard concepts. This step also involves conversion of the source vocabulary structure into vocabulary-agnostic staging data structure
3. QA/QC Part I (Quality Assessment): Initial quality checks focused on staging tables, including source data completeness, format validation, and data type conformity before integration into OMOP tables
4. Generic Update (Vocabulary Integration): Integration of staged vocabularies into OMOP tables, establishing concept IDs and relationships
5. QA/QC Part II (Final Validation): Validation for integrated vocabulary that combines automated quality checks for data integrity and change tracking, complemented by systematic manual review focusing on concept changes and their relationships

### Stage 4: Publication

Following successful validation, we collaborated with the OHDSI Vocabulary Team[13] to address feedback and refine mappings. We submitted the outputs of our pipeline as the Community Contribution inputs, including ‘sources.edi_data’, ‘concept_stage’, ‘concept_synonym_stage’, ‘concept_relationship_stage’, and ‘concept_relationship_manual’ to receive final feedback. Additionally, to ensure code transparency and reproducibility, we committed the open-source code (SQL queries) used in the Community Contribution Pipeline process as issues to the OHDSI GitHub Vocabulary repository[14]. The integrated vocabulary that passed all requirements was published on Athena[15], the official OHDSI Vocabularies website.

## RESULT

Building upon our previous work, which used only data from October 2019[5], the current study collected data spanning from 2000 to 2023 and mapped them with standard concepts following the mapping guidelines.

Integration Coverage: Of the total 620,642 EDI codes collected from November 2000 to May 2024, we successfully integrated 437,807 codes (70.5%) into the OHDSI Vocabularies system with 376,111 standard concepts. Table 2 presents the domain-specific integration coverage, showing the proportion of collected codes that were successfully integrated.

**Table 2.** Domain-specific Integration Coverage of Korean EDI Codes.

| Domain | Collected Codes | Integrated Codes | Integration Coverage (%) |
| --- | --- | --- | --- |
| Total | 620,642 | 376,111 | 60.6 |
| Drug | 65,982 | 62,038 | 94.0 |
| Device | 45,133 | 35,850 | 79.4 |
| Procedure | 444,022 | 276,559 | 62.3 |
| Measurement | 65,509 | 1,664 | 2.5 |

Mapping Coverage: Table 3 presents a detailed comparison between previous and current code integration efforts in terms of mapping to standard concepts. Of the 437,807 integrated EDI codes, 376,111 (85.9%) were successfully mapped to standard concepts, compared to 0.5% (1,606/313,431) in previous attempt.

**Table 3.** Comparison of Mapping Coverage between Previous and Current Studies.

| Domain | Before |  |  | After |  |  |
| --- | --- | --- | --- | --- | --- | --- |
|  | Source Code | Source Mapped | Mapping Coverage (%) | Source Code | Source Mapped | Mapping Coverage (%) |
| Total | 313,431 | 1,606 | 0.5 | 437,807 | 376,111 | 85.9 |
| Drug | 23,231 | 0 | 0.0 | 62,064 | 62,038 | 99.9 |
| Device | 19,813 | 0 | 0.0 | 36,196 | 35,850 | 99.0 |
| Procedure | 249,785 | 1,606 | 0.6 | 318,945 | 276,559 | 86.7 |
| Measurement | 20,602 | 0 | 0.0 | 20,602 | 1,664 | 8.1 |

The integrated vocabulary passed all Community Contribution Pipeline requirements, including concept relationship consistency, domain classification accuracy, referential integrity validation, and terminology standard compliance. The final vocabulary was published in September 2024 on Athena [15], the official OHDSI vocabulary repository, enabling its use across the OHDSI network.

## DISCUSSION

Our experience with integrating Korean EDI code demonstrates potential of local vocabulary standardization. By following the methodology outlined in this tutorial, we achieved an overall integration coverage of 70.5% (437,807/620,642) of all collected EDI codes, with 85.9% (376,111/437,807) of these integrated codes successfully mapped to standard concepts. This is a substantial improvement compared to the previous 0.5% (1,606/313,431) mapping coverage.

While the overall integration and mapping coverage showed substantial improvement, challenges remain in specific domains. The particularly low coverage in the Measurement domain (2.5% integration, 8.1% mapping of integrated codes) warrants further discussion. This limitation stems from the high complexity and specificity of laboratory tests in the Korean healthcare system that often lack direct equivalents in international terminologies. To address these challenges, future work should focus on developing automated approaches to improve coverage while maintaining accuracy.

The successful integration of Korean EDI codes demonstrates that local vocabularies with different structures, languages, and update patterns can be effectively incorporated into standardized data models. This approach contributes to the global OHDSI mission of generating reliable evidence from observational health data through international collaboration.

## Data Availability

All data produced in the present work are contained in the manuscript

## ACKNOWLEGMENT

This work was supported by the National Research Foundation of Korea (NRF) grant funded by the Korea government (MSIT) (RS-2024-00341426).

