## Supplemental information for "Integrating Local Vocabulary into OMOP CDM: A Step-by-Step Tutorial"

Oct 24, 2024

### Korean EDI Updates in ATHENA

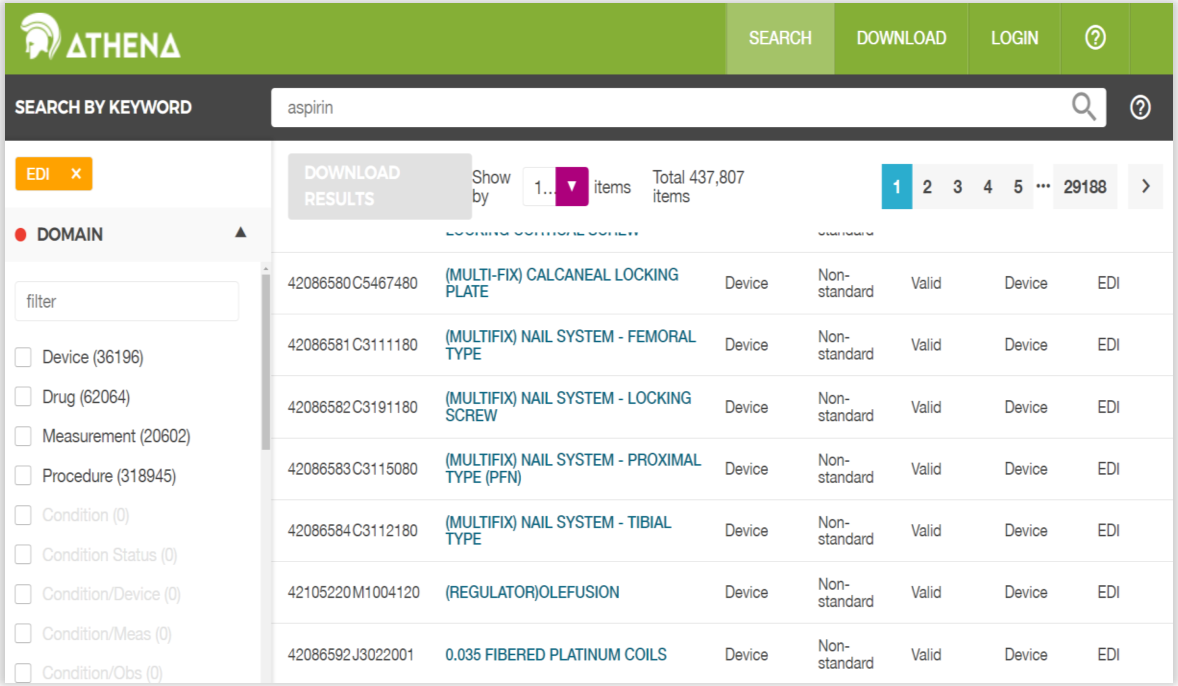


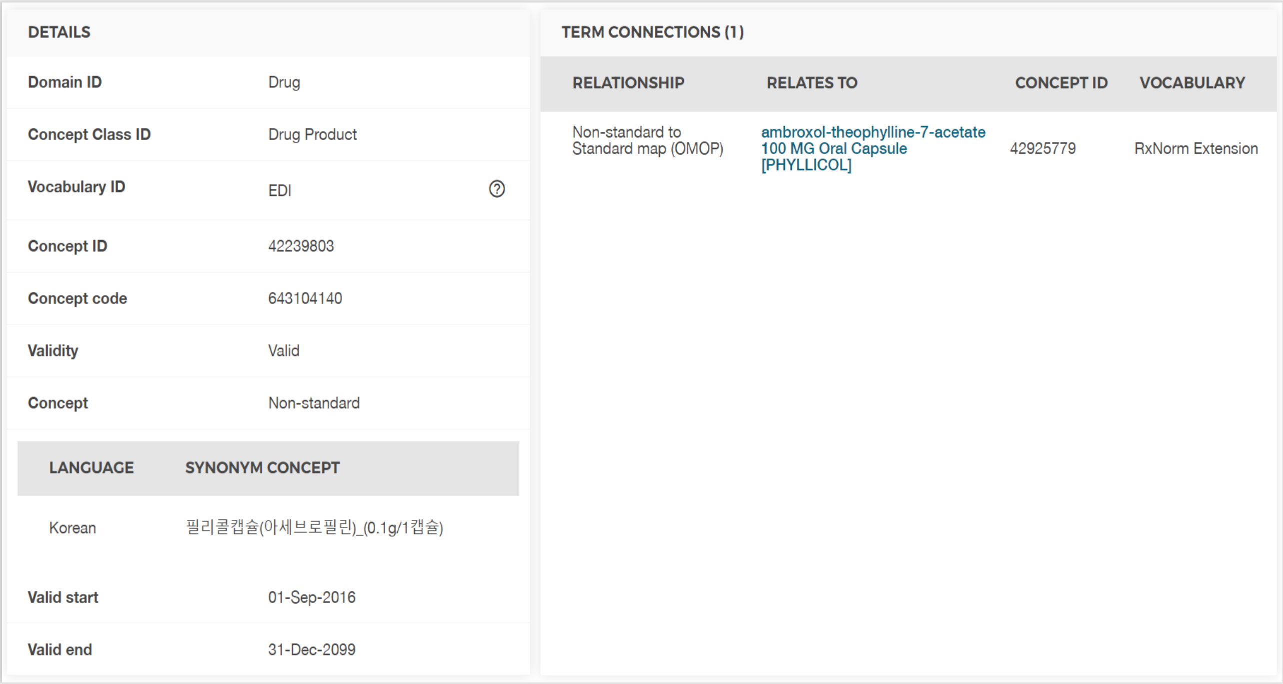


We integrated an updated version of the Korean EDI in September 2024

### Topics

- Introduce the **Korean EDI** Vocabulary
- **Process** for Data Integration in ATHENA
    - Create a Semi-automated process : **SYNC package**
    - Mapping to Standard Concepts
    - Community Contribution Process

### Introduction of EDI

- **EDI (Electronic Data Interchange) is a widespread code system for the reimbursement or claim data in Korea.**

- EDI concepts are divided into **drugs**, **devices**, and **medical services**.

- EDI is developed and maintained by **HIRA** (Health Insurance Review & Assessment Service).

- EDI is **updated every month** and is a **longitudinal data**.

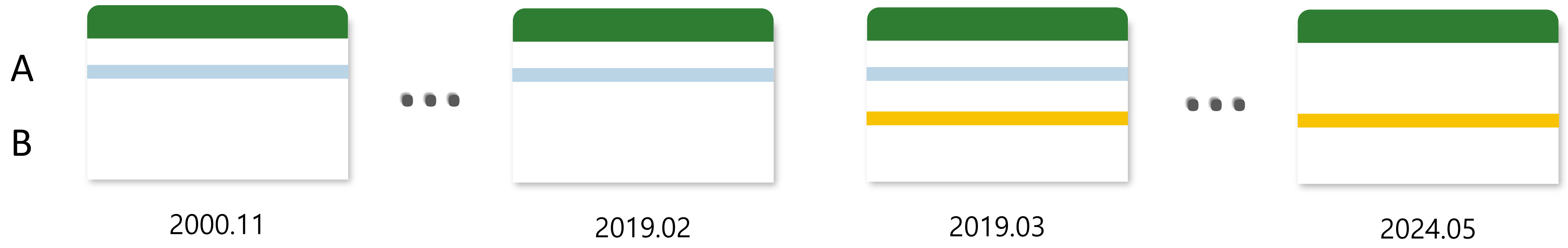


Example of EDI data from HIRA (each line represents one EDI code)

### Introduction of EDI

- Despite widespread adoption in Korean EHR systems, **limitations** still persist.
  - Validity dates are not recorded in the official monthly announcement.
  - There are expired or replaced EDI code and outdated EDI can be assigned to new concepts.
  - EDI has duplicated identifiers due to the lack of a unified encoding system across domains.

#### OUR GOAL WAS …

To enhance EDI vocabulary for a controlled and standardized vocabulary system

- For this purpose, we incorporated the EDI into OMOP vocabulary **using a semi-automated process**.

### SYNC package

- SYNC package is a **semi-automated process** we made.
- This package makes EDI vocabulary as a Source Concepts using OMOP structure.
- There are **four main steps** to incorporate EDI into OHDSI standardized vocabulary.

1 **Scrap: Crawl EDI data from HIRA website**

2 **Yield: Categorize codes into domains (Device, Drug, Procedure, Measurement)**

3 **Nest: Built a vertical hierarchy between EDI concepts**

4 **Convert: Added an English definition for each EDI concept using Google Translation**

### SYNC package

#### 1 Crawl EDI data from HIRA website

- Through web crawling, all necessary EDI data from the HIRA website is downloaded
- There are three types of data provided by HIRA: Device, Drug, Medical Service
- We have collected all EDI data from November 2000 to May 2024

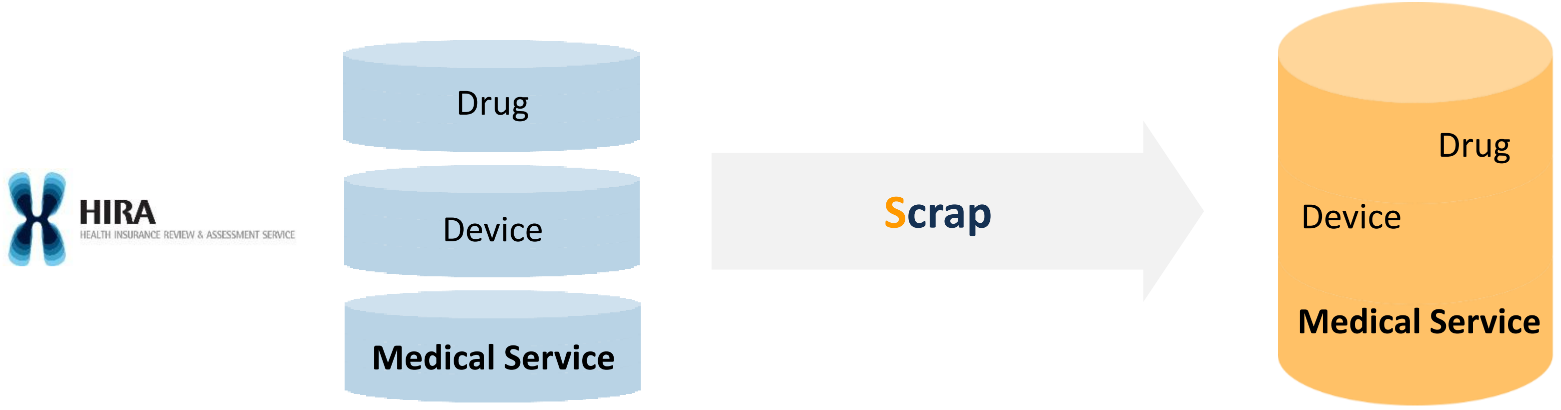

### SYNC package

#### 2 Categorize codes into domains (Device, Drug, Procedure, Measurement)

- As mentioned, EDI concepts are divided into **Drugs**, **Devices**, and **Medical Services**.
- The scope of **Medical Services is too broad** for the OHDSI standardized vocabularies.
- So, we subclassified Medical Services into **Procedures** and **Measurements** to match the OHDSI domains.

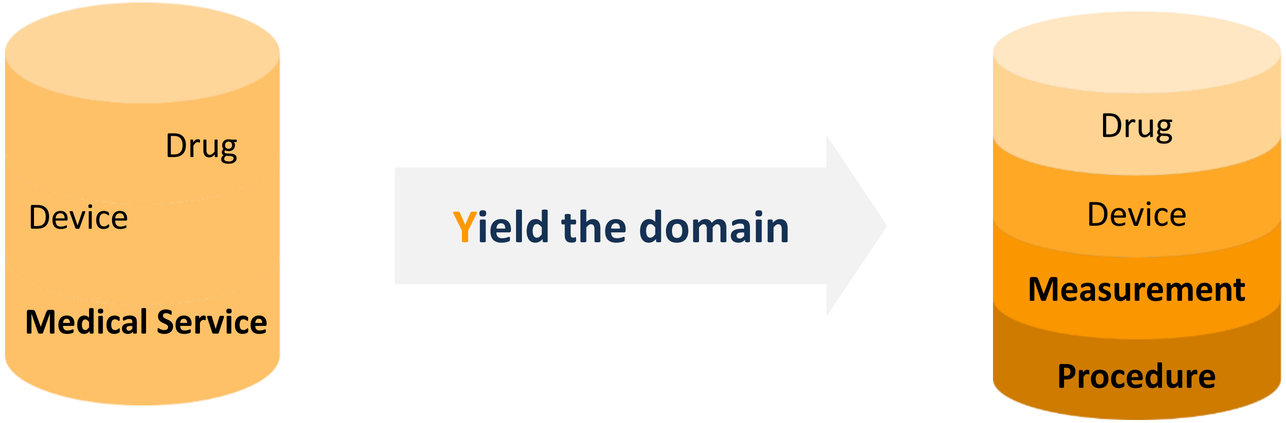

### SYNC package

#### 2 Categorize codes into domains (Device, Drug, Procedure, Measurement)

##### +) Assigned three attributes for each EDI concept

Attributes: Valid start date, Valid end date, Invalid reason

- **Valid start date, Valid end date**
  - When an EDI concept is newly registered or deprecated, the term's date is updated or expired.
- **Invalid reason**
  - If a concept is valid ⟶ invalid reason : NULL
  - If a concept is replaced by another concept or deleted ⟶ invalid reason : U or D

| Concept code | Valid start date | Valid end date | Invalid reason |
|---|---|---|---|
| A29506361 | 2008-04-01 | 2099-12-31 | NULL |

newly registered ⟶

| Concept code | Valid start date | Valid end date | Invalid reason |
|---|---|---|---|
| A29506361 | 2008-04-01 | **2010-01-31** | **U** |
| 670600010 | **2010-02-01** | 2099-12-31 | NULL |

### SYNC package

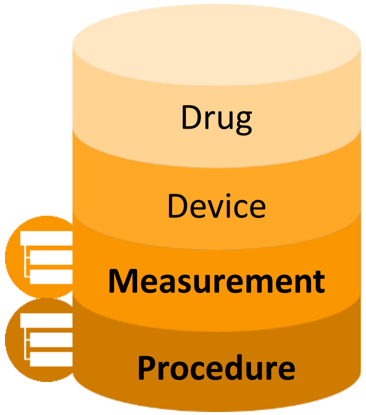


#### 3 Built a vertical hierarchy between EDI concepts

- We built a formal vertical hierarchy for EDI concepts as ICD-9 or ICD-10 code system.

- The **first five digits** of the EDI code in the medical service domain (procedure, measurement) **represent the ancestor terms** for longer descendent EDI code.

| Concept code | Concept name | Ancestor concept code |
|---|---|---|
| HC281 | Whole Body Scan | |
| HC281006 | Whole Body Scan, Nuclear Medicine and other physician reading | HC281 |
| HC281300 | Whole Body Scan, Under 8 years old | HC281 |
| HC281306 | Whole Body Scan, Under 8 years old, read by nuclear medicine physician | HC281 |
| HC281600 | Whole Body Scan, Under 72 months | HC281 |
| HC281606 | Whole Body Scan, Under 72 months, Nuclear Medicine physician reading | HC281 |

**ancestor** (HC281)

**descendent** (HC281006 – HC281606)

### SYNC package

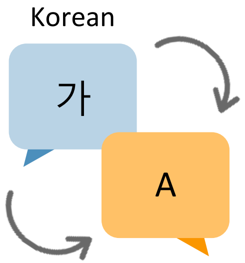


#### 4 Added an English definition for each EDI concept using Google Translation

- We have to add an English definition for each EDI term.
- We utilized **Google Cloud Translation API** for the initial translation.
- Inaccurately translated words underwent **review by nurses** and were retranslated.

| Korean definition | Using Google Translation API | Using Google Translation API With glossary |
|---|---|---|
| M핵산증폭-정량그룹1_B형 감염바이러스 [중합효소연쇄반응교잡반응법] | Nucleic acid amplification-quantitative group 1 hepatitis B virus [polymerase chain reaction hybridization method] | Nucleic acid amplification-quantitative group 1_HBV [PCR-Hybridization] |
| 단기사용담관용튜브・카테터 | Short-term use bile duct tube and catheter | Cahteter, bile duct short-term use |

### SYNC package

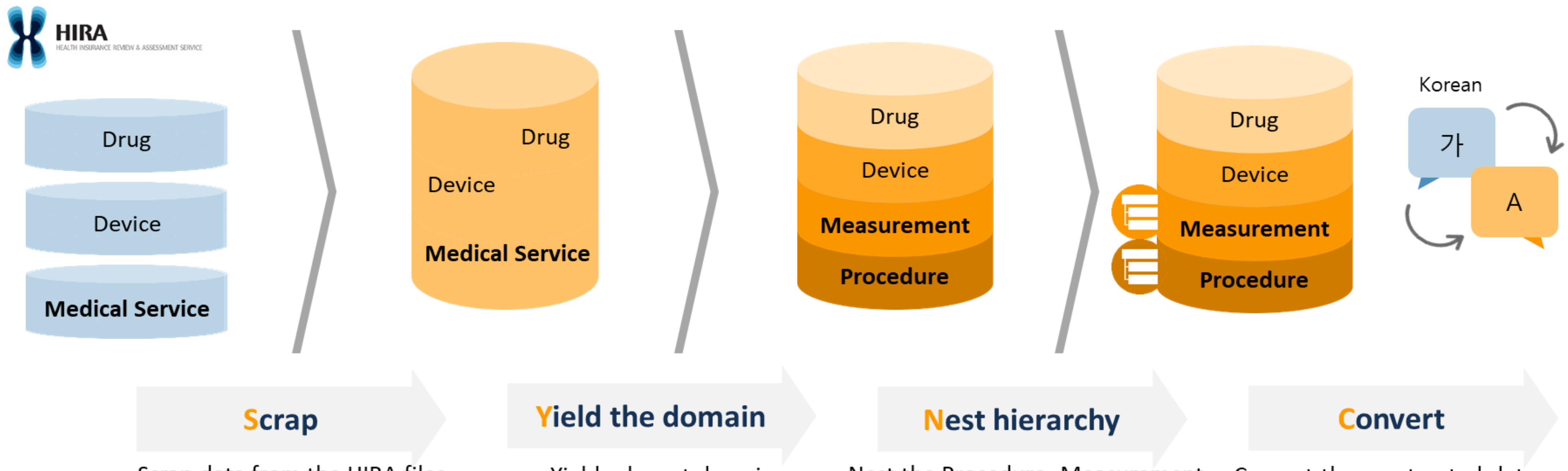


Scrap data from the HIRA files

Yield relevant domain classification

Nest the Procedure, Measurement domains into a hierarchical structure

Convert the constructed data Korean to English

### SYNC package

| concept_ | concept_name | concept_synonym | domain_id | vocab | concept_class_id | valid_start_ | valid_end_ | inv | ances | mater | comp | dosag | dosag | sanju | name |
|---|---|---|---|---|---|---|---|---|---|---|---|---|---|---|---|
| O2055 | Thrombectomy(Artery),Neck | 혈전제거술(동맥-경부) | Procedure | EDI | Proc Hierarchy | 2008-01-01 | 2099-12-31 | | | | | | | | |
| O2055010 | Thrombectomy(Artery),Neck,Nighttime | 혈전제거술(동맥-경부),야간 | Procedure | EDI | Procedure | 2008-01-01 | 2099-12-31 | | O2055 | | | | | 야간 | |
| O2055020 | Thrombectomy(Artery),Neck,emergency | 혈전제거술(동맥-경부),응급 | Procedure | EDI | Procedure | 2016-01-01 | 2023-12-31 | D | O2055 | | | | | 응급 | |
| D3030 | 1.5-Anhydro-D-Glucitol | 1.5-Anhydro-D-Glucitol[화학빈 | Measurement | EDI | Proc Hierarchy | 2018-01-01 | 2099-12-31 | | | | | | | | |
| D3030001 | 1.5-Anhydro-D-Glucitol,Diagnostic and laboratory te | 1.5-Anhydro-D-Glucitol[화학빈 | Measurement | EDI | Proc Hierarchy | 2018-01-01 | 2099-12-31 | | D3030 | | | | | 진단검사 질가산(4%) | |
| D3030002 | 1.5-Anhydro-D-Glucitol,Diagnostic and laboratory te | 1.5-Anhydro-D-Glucitol[화학빈 | Measurement | EDI | Proc Hierarchy | 2018-01-01 | 2099-12-31 | | D3030 | | | | | 진단검사 질가산(3%) | |
| 646902600 | acetaminophen(encapsulated) 0.65g | 타이레놀8시간이알서방정( | Drug | EDI | Drug Product | 2022-07-01 | 2022-12-31 | U | 101430ATR | | (주)한국 | 1 | 정 | | |
| 672300240 | acetaminophen(encapsulated) 0.65g | 타이레놀8시간이알서방정( | Drug | EDI | Drug Product | 2023-01-01 | 2099-12-31 | | 101430ATR | | 한국존[illegible] | 1 | 정 | | |
| G2201003 | COREVALVE EVOLUT SYSTEM | | Device | EDI | Device | 2015-06-01 | 2099-12-31 | | | PORCINE | 메드트로[illegible] | 전규격 | 1KIT | | |
| G2201002 | EDWARDS SAPIEN 3 AND SAPIEN 3 ULTRA TRANSCA | | Device | EDI | Device | 2015-06-01 | 2099-12-31 | | | BOVINE P | 에드워즈 | 전규격 | 1KIT | | |

- After completing all processes in the SYNC package, we generate a merged vocabulary tables as shown.
- These tables contain comprehensive information(material, company names, dosage) about each EDI code.

### Mapping to Standard Concepts

- EDI concepts were mapped to standard concepts following the OHDSI-Korea community guidelines
- Every concepts should be **standard** based on the ATHENA

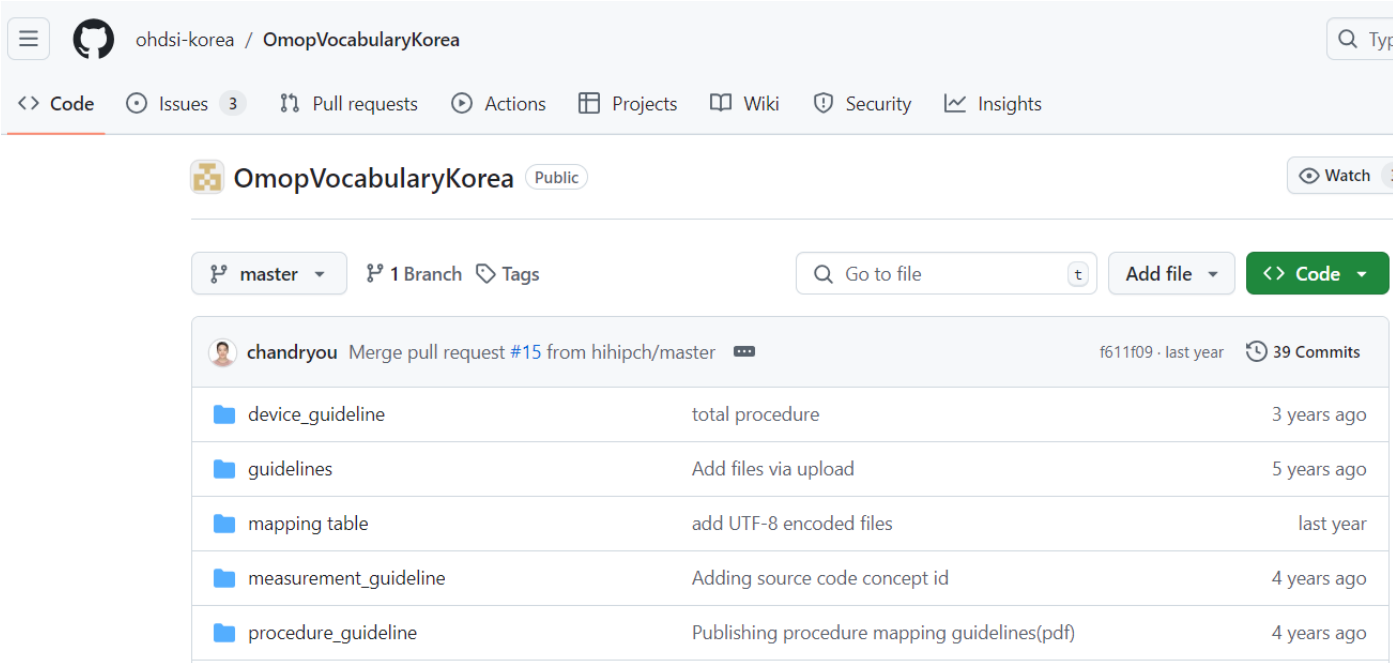


< Priority of Standard Concepts in Mapping >

| | Non-standard Concepts | Standard Concepts |
|---|---|---|
| Procedure | EDI | SNOMED |
| Measurement | EDI | 1) LOINC<br>2) SNOMED |
| Drug | EDI | RxNorm<br>RxNorm Extension |
| Device | EDI | SNOMED |

### 1. Procedure Mapping

1. [http://athena.ohdsi.org/](http://athena.ohdsi.org/)
2. Filter
   - Domain: Procedure
   - Standard Concept: Standard
   - Vocabulary: SNOMED

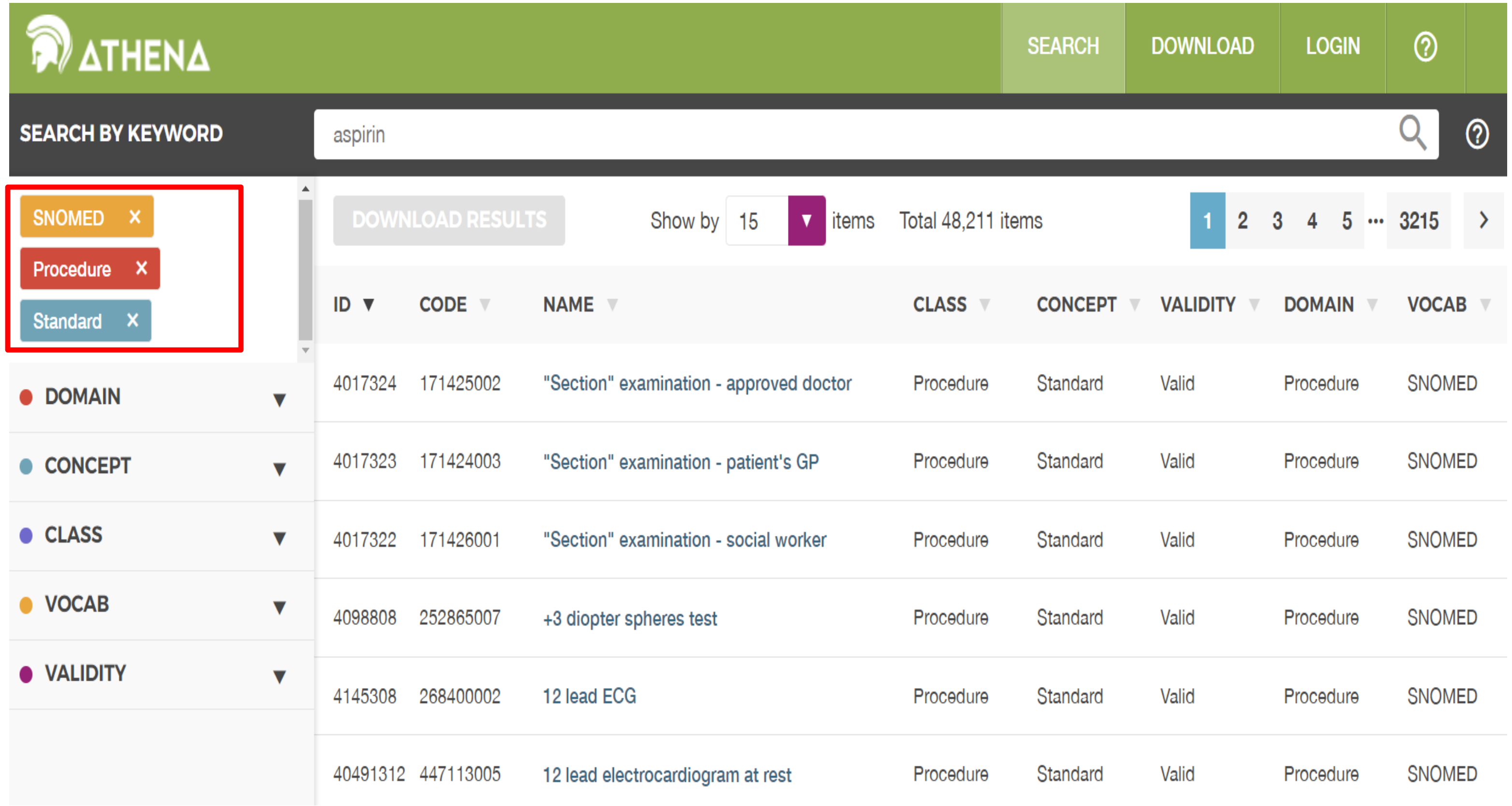

### 1. Procedure Mapping

If you want to map ‘Hemiarthroplasty of hip’

1. Search your procedure name in ATHENA

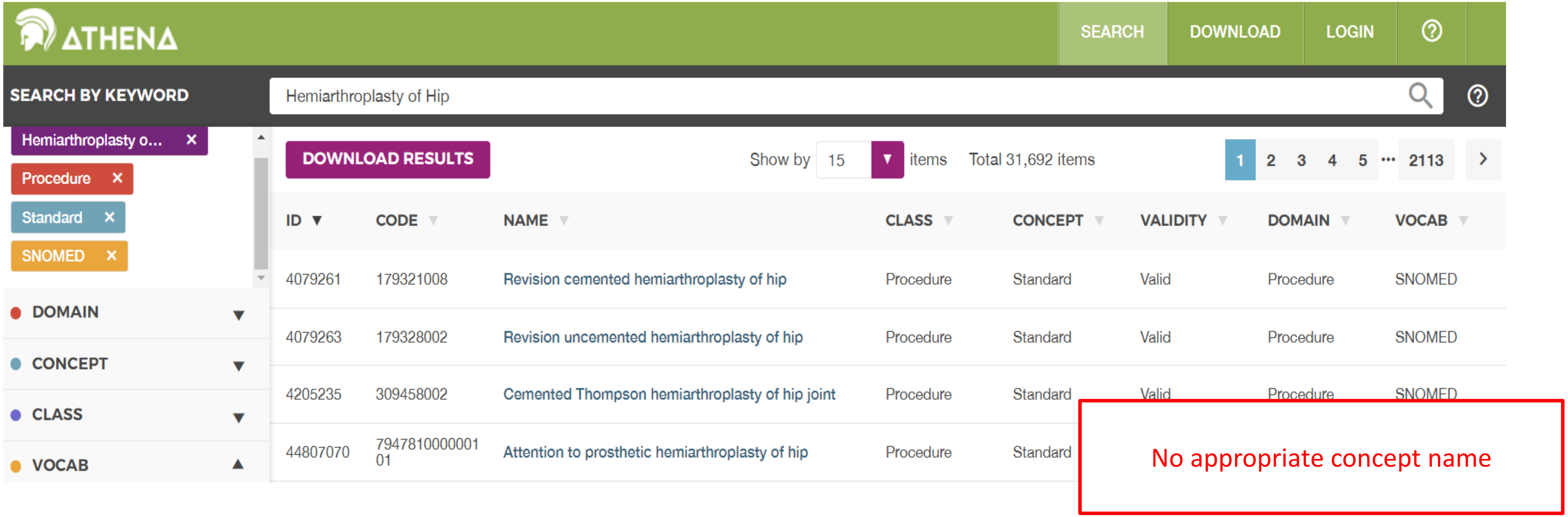

### 1. Procedure Mapping

If you want to map ‘Hemiarthroplasty of hip’

2. Modify Search Terms and Re-search

   e.g., Hemiarthroplasty → partial joint replacement

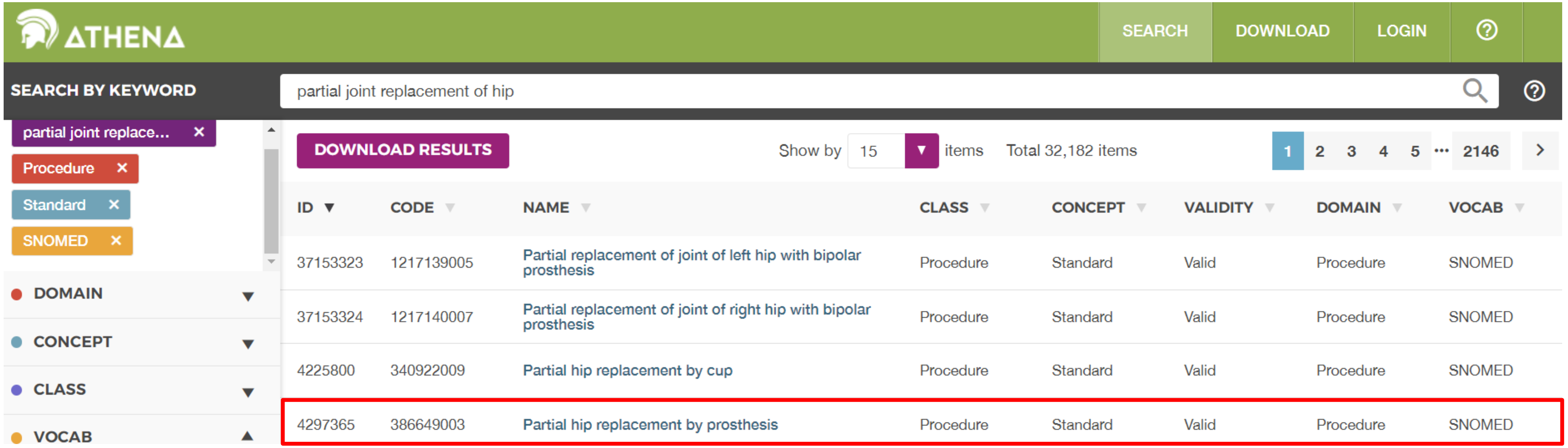

### 1. Procedure Mapping

If you want to map 'Hemiarthroplasty of hip'

3. Move up to broader concepts, and check subsumes for better matches

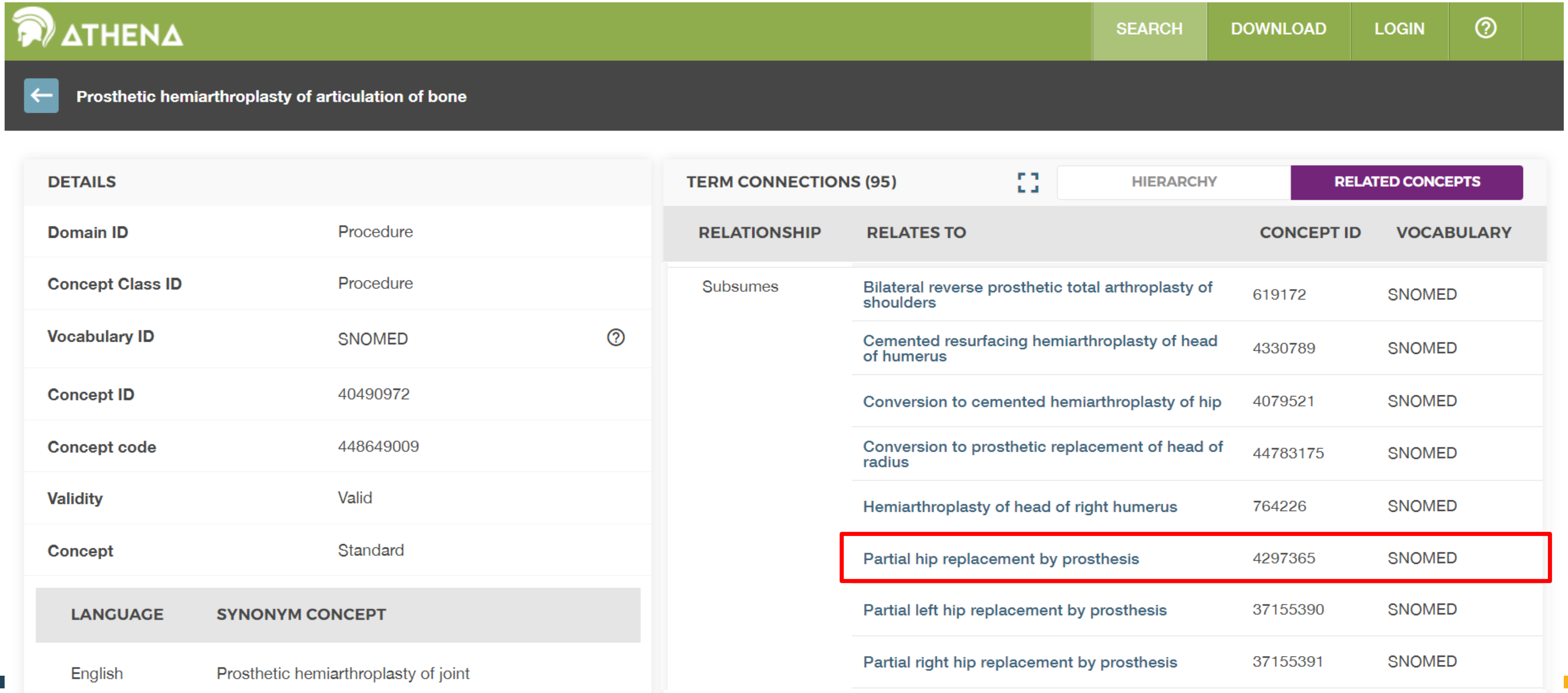

### 2. Measurement Mapping

1. [http://athena.ohdsi.org/](http://athena.ohdsi.org/)
2. Filter
   - Domain: Measurement
   - Standard Concept: Standard
   - Vocabulary: LOINC(mostly)

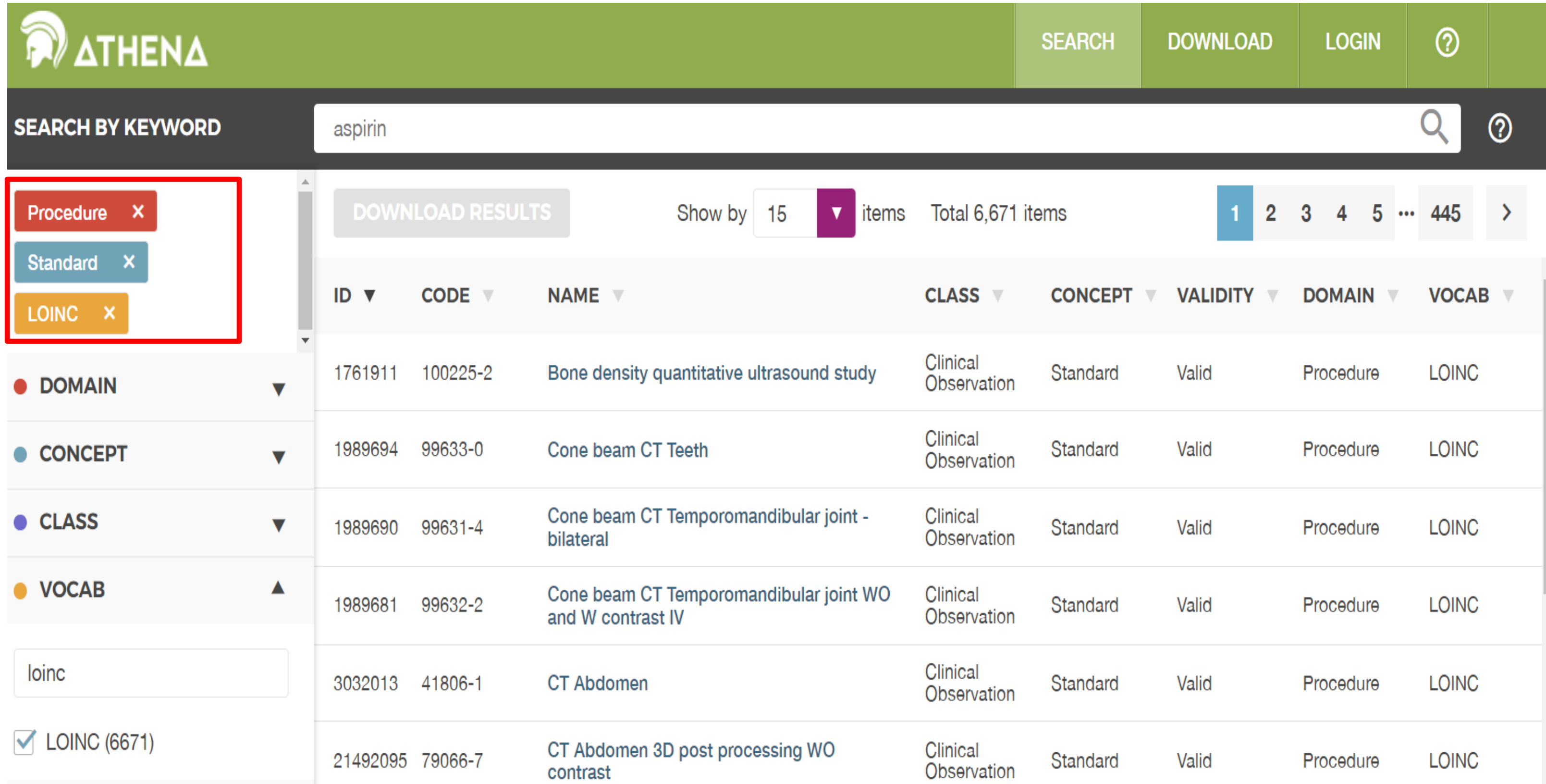

### Measurement Mapping

- Search with the unit and specimen type

1. Unit: mg/dL, Specimen: serum
    - mg/dL → [mass/volume]
    - Search for ‘ [mass/volume] in serum’ in ATHENA

2. Unit: g/dL, Specimen: whole blood
    - g/dL → [mass/volume]
    - Search for ‘[mass/volume] in whole blood’ in ATHENA

| | | |
|---|---|---|
| **dL** | 1/10 L | Volume |
| **mL** | 1/1,000 L | Volume |
| **mg** | 1/1,000 g | Mass |
| **µg** | 1/1,000,000 g | Mass |
| **Ng** | 1/1,000,000,000 g | Mass |
| **Pg** | 1/1,000,000,000,000 g | Mass |
| **U** | Standardized Units for Each Test | Units<br>Enzymatic activity |
| **mIU** | 1/1,000 U | Units |
| **µU** | 1/1,000,000 U | Units |
| **mol** | Quantity of Atoms and Molecules | Moles |
| **mmol** | 1/1,000 mol | Moles |
| **µmol** | 1/1,000,000 mol | Moles |
| **nmol** | 1/1,000,000,000 mol | Moles |

### 2. Measurement Mapping

- When it's difficult to choose between similar concepts, Go to https://loinc.org/

LOINC from Regenstrief

Learn | Content | Downloads | Community

HOME ▸ CONTENT

#### Download LOINC

##### Other ways to access LOINC content

A LOINC login is required to access each of these resources. You can create one for free.

**SearchLOINC**
This web-based tool allows you to find and export LOINC codes using an intuitive interface.

**LOINC API using HL7® FHIR®**
Access LOINC content programmatically using the HL7's popular FHIR standard.

**RELMA®**
Download and install this program for Windows to search for LOINC terms and map them to your local codes.

**Archive of past LOINC releases**
A complete archive of previous releases is available for your review.

**SearchLOINC**
Find LOINC concepts in your browser with this powerful web app. A free LOINC login is required.

**Hierarchy Browser**
Instead of searching, peruse LOINC codes via numerous hierarchies. A free LOINC login is required.

**Community Forum**
Ask questions and chime in on conversations with your fellow LOINCers.

**Request a LOINC**
Cannot find an appropriate LOINC concept? Follow instructions for requesting a new code.

**Submission Queue**
See all the new concepts we are working on now, including any submissions you may have made.

LOINC from Regenstrief

Learn | Content | Downloads | Community

HOME ▸ CONTENT

#### LOINC Mapping Guides

**LOINC Mapping Guide - Allergy**
Published September 2022.
DOWNLOAD Version 1.0

**LOINC Mapping Guide - Cell Markers**
Published September 2022.
DOWNLOAD Version 1.0

**LOINC Mapping Guide - Chemistry**
Published September 2022.
DOWNLOAD Version 1.0

**LOINC Mapping Guide - Drug and Toxicology**
Published September 2022.
DOWNLOAD Version 1.0

**LOINC Mapping Guide - Hematology and Serology**
Published September 2022.
DOWNLOAD Version 1.0

**LOINC Mapping Guide - Molecular Pathology**
Published September 2022.
DOWNLOAD Version 1.0

**Guide for Using LOINC Microbiology Terms**
Originally published August 2018.
Version 1.1 released in August 2019.
DOWNLOAD Version 1.1

### 2. Measurement Mapping

1. RELMA
   - Detailed information: component, property, timing, system, scale, method
   - Concept ranking
     - Higher ranks prioritized
     - Indicates common usage

2. Hierarchy Browser
   - Visualize concept relationships

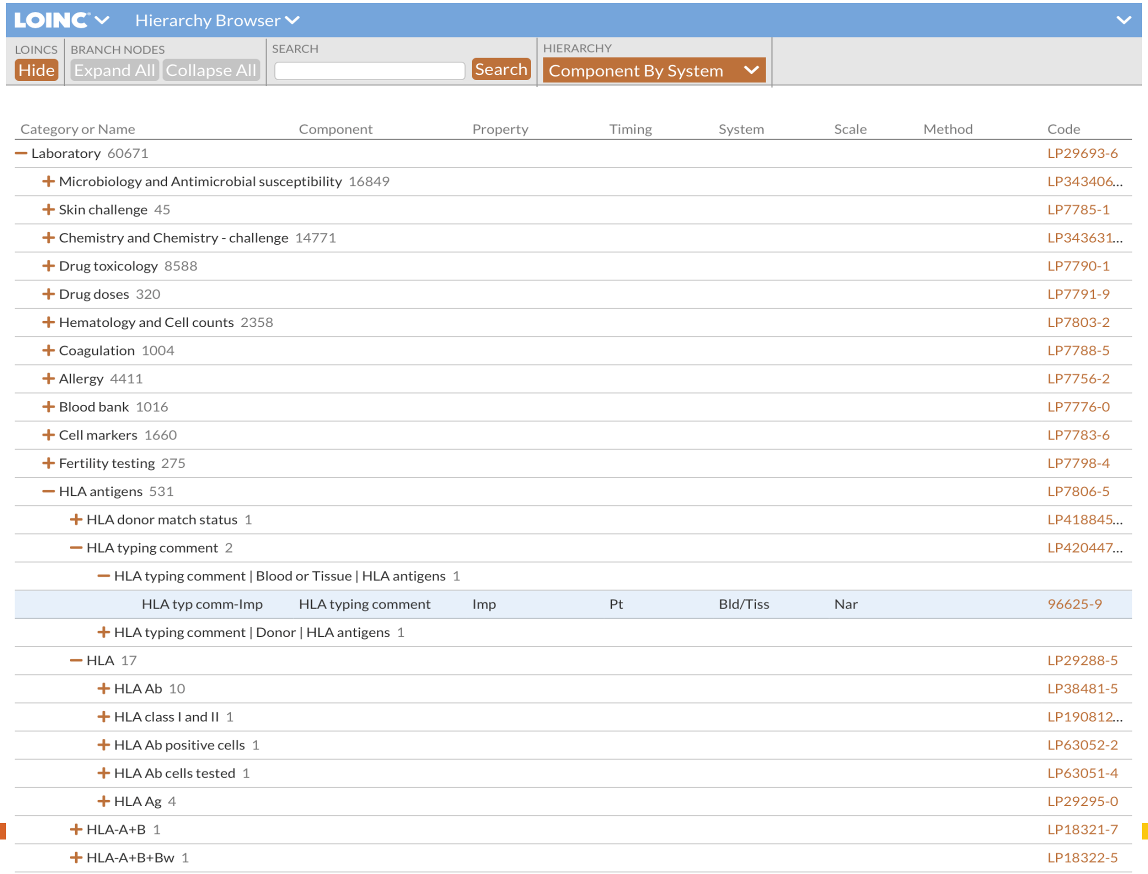

### 3. Drug Mapping

- Mapping to **RxNorm** or **RxNorm Extension**
- **Mapping Priority**

1. Ingredient + Dosage + Dosage Form + [Drug Name] + by Company Name

   Acetaminophen + 160 mg + chewable tablet + [TAIFEN] + by Youngpoong — Concept id: **2071554**

2. Ingredient + Dosage + Dosage Form + [Drug Name]

   Acetaminophen + 160 mg + chewable tablet + [TAIFEN] — Concept id: **2071553**

3. Ingredient + Dosage + Dosage Form

   Acetaminophen + 160 mg + chewable tablet — Concept id: **19079924**

4. Ingredient + Dosage Form

   Acetaminophen + chewable tablet — Concept id: **40005631**

5. Ingredient only

   Acetaminophen — Concept id: **1125315**

### 3. Drug Mapping

- We need to distinguish between **Injection** and **Injectable solution**
  - MG → Injection
  - MG/ML → Injectable solution

- **Injection**
  - A pharmaceutical preparation that is administered directly into the body through the skin or mucous membranes, available as solutions, suspensions, emulsions, or as powder forms that need to be dissolved or suspended before use
  - **E.g. Powder injections in MG, G forms**
- **Injectable Solution**
  - A ready-to-use injection containing one or more drugs in a suitable solvent or mixture for injection
  - **E.g. Injections in MG/ML forms**

### 4. Device Mapping

- Mapping to **SNOMED**
- Utilize company name or materials
- (Only for EDI) Recommended to refer to the ‘Medical Device Insurance Guide Map’ published by HIRA

### Community Contribution Process

- For the integration, you need to adhere to the **Community Contribution pipeline**.
- If you are interested in loading the vocabulary, please contact the Vocabulary Team.

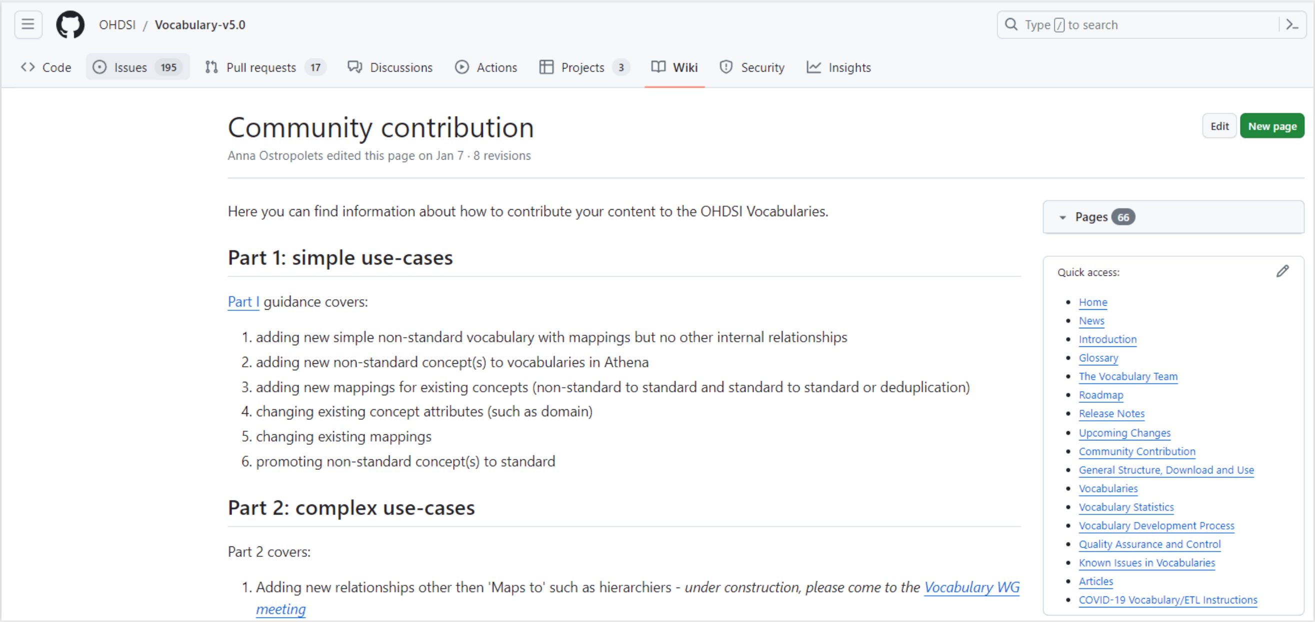
OHDSI / Vocabulary-v5.0

Type / to search

Code | Issues 195 | Pull requests 17 | Discussions | Actions | Projects 3 | Wiki | Security | Insights

Edit | New page

Community contribution

Anna Ostropolets edited this page on Jan 7 · 8 revisions

Here you can find information about how to contribute your content to the OHDSI Vocabularies.

Part 1: simple use-cases

Part I guidance covers:

1. adding new simple non-standard vocabulary with mappings but no other internal relationships
2. adding new non-standard concept(s) to vocabularies in Athena
3. adding new mappings for existing concepts (non-standard to standard and standard to standard or deduplication)
4. changing existing concept attributes (such as domain)
5. changing existing mappings
6. promoting non-standard concept(s) to standard

Part 2: complex use-cases

Part 2 covers:

1. Adding new relationships other then 'Maps to' such as hierarchiers - *under construction, please come to the Vocabulary WG meeting*

Pages 66

Quick access:

- Home
- News
- Introduction
- Glossary
- The Vocabulary Team
- Roadmap
- Release Notes
- Upcoming Changes
- Community Contribution
- General Structure, Download and Use
- Vocabularies
- Vocabulary Statistics
- Vocabulary Development Process
- Quality Assurance and Control
- Known Issues in Vocabularies
- Articles
- COVID-19 Vocabulary/ETL Instructions

**Contact the Vocabulary Team**

**or**

**Vocabulary WG meeting.**

### Community Contribution Process

- **Prerequisites**
  - Preparation Schemas : *sources*, *dev_xyz*, *devv5*
  - Copies of tables, fully indexed (downloaded from Athena and put into Schema *dev_xyz*, *devv5*)

- **Process**
  1. Run *load_stage.sql* in the *dev_xyz* schema
  2. QA/QC part 1
  3. Generic update
  4. QA/QC part 2 : semi-automatic process

### Community Contribution Process

- **Prerequisites**
  - Preparation Schemas : *sources*, *dev_xyz*, *devv5*
  - Copies of tables, fully indexed (downloaded from Athena and put into Schema *dev_xyz*, *devv5*)

- **Process**
  1. Run *load_stage.sql* in the *dev_xyz* schema
  2. QA/QC part 1
  3. Generic update
  4. QA/QC part 2 : semi-automatic process

### Community Contribution Process

- **Prerequisites**
  - **Preparation Schemas : *sources, dev_xyz, devv5***
  - Copies of tables, fully indexed (downloaded from Athena and put into Schema *dev_xyz*, *devv5*)

**Schema *sources***

- Put your **source vocabulary**

**Schema *dev_xyz***

- Set as **Working directory**
- Run *DevV5_DDL.sql* to create empty tables
- Put copy of vocabularies downloaded from Athena

**Schema *devv5***

- **Reference Schema** of vocabularies from Athena
- Run DevV5_DDL.sql to create empty tables
- Put copy of vocabularies downloaded from Athena

### Community Contribution Process

- **Prerequisites**
  - Preparation Schemas : *sources, dev_xyz, devv5*
  - **Copies of tables, fully indexed (downloaded from Athena and put into Schema *dev_xyz, devv5*)**

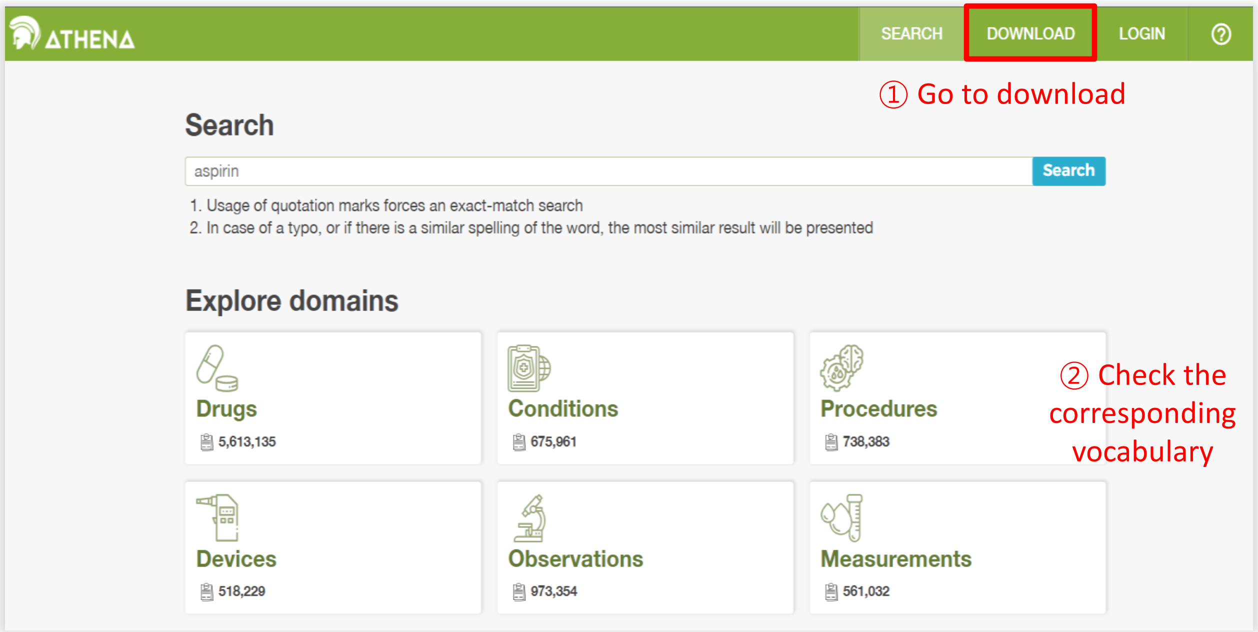


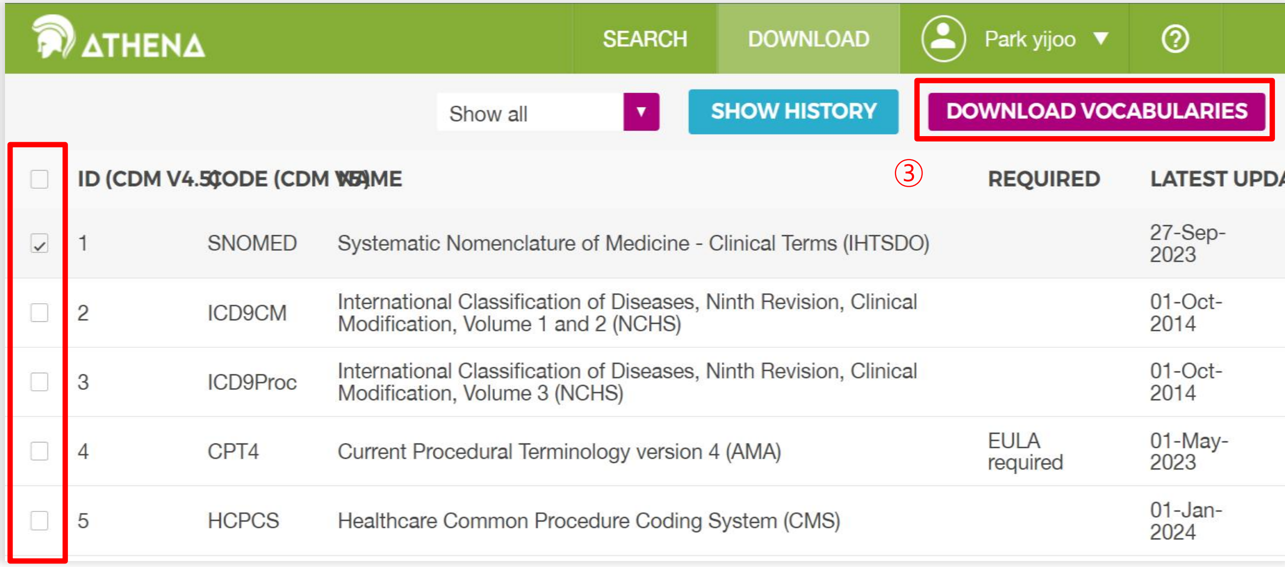

### Community Contribution Process

- **Prerequisites**
  - Preparation Schemas : *sources*, *dev_xyz*, *devv5*
  - **Copies of tables, fully indexed (downloaded from Athena and put into Schema *dev_xyz*, *devv5*)**

Vocabulary-v5.0 / working / DevV5_DDL.sql

Code Blame 314 lines (288 loc) · 14.1 KB

```
--Main DDL

DROP TABLE IF EXISTS concept CASCADE;
CREATE TABLE concept (
            concept_id int4 NOT NULL,
            concept_name VARCHAR (255) NOT NULL,
            domain_id VARCHAR (20) NOT NULL,
            vocabulary_id VARCHAR (20) NOT NULL,
            concept_class_id VARCHAR (20) NOT NULL,
            standard_concept VARCHAR (1),
            concept_code VARCHAR (50) NOT NULL,
            valid_start_date DATE NOT NULL,
            valid_end_date DATE NOT NULL,
            invalid_reason VARCHAR (1)
);
```

④ Run --Main DDL in postgreSQL

Tables (21)

- concept
- concept_ancestor
- concept_class
- concept_class_conversion
- concept_manual
- concept_relationship
- concept_relationship_manual
- concept_relationship_stage
- concept_stage
- concept_synonym
- concept_synonym_manual
- concept_synonym_stage
- domain
- drug_strength
- drug_strength_stage
- pack_content
- pack_content_stage
- relationship
- relationship_conversion
- vocabulary
- vocabulary_conversion

⑤ Upload the corresponding tables from the vocabulary bundle in Athena

Vocabulary-v5.0 / working / DevV5_DDL.sql

Code Blame 314 lines (288 loc) · 14.1 KB

```
--Create PKs
ALTER TABLE concept ADD CONSTRAINT xpk_concept PRIMARY KEY (concept_id);
ALTER TABLE vocabulary ADD CONSTRAINT xpk_vocabulary PRIMARY KEY (vocabulary_id);
ALTER TABLE domain ADD CONSTRAINT xpk_domain PRIMARY KEY (domain_id);
ALTER TABLE concept_class ADD CONSTRAINT xpk_concept_class PRIMARY KEY (concept_class_id)
ALTER TABLE concept_relationship ADD CONSTRAINT xpk_concept_relationship PRIMARY KEY (con
ALTER TABLE relationship ADD CONSTRAINT xpk_relationship PRIMARY KEY (relationship_id);
ALTER TABLE concept_ancestor ADD CONSTRAINT xpkconcept_ancestor PRIMARY KEY (ancestor_con
ALTER TABLE drug_strength ADD CONSTRAINT xpk_drug_strength PRIMARY KEY (drug_concept_id,

--Create external keys
ALTER TABLE concept ADD CONSTRAINT fpk_concept_domain FOREIGN KEY (domain_id) REFERENCES
ALTER TABLE concept ADD CONSTRAINT fpk_concept_class FOREIGN KEY (concept_class_id) REFER
ALTER TABLE concept ADD CONSTRAINT fpk_concept_vocabulary FOREIGN KEY (vocabulary_id) REF
```

⑥ Create PKs, external keys, indexes and checks

https://github.com/OHDSI/Vocabulary-v5.0/blob/bd54f495932acbe529d4534eae21195aa6455759/working/DevV5_DDL.sql

### Community Contribution Process

- **Prerequisites**
  - Preparation Schemas : *sources*, *dev_xyz*, *devv5*
  - Copies of tables, fully indexed (downloaded from Athena and put into Schema *dev_xyz*, *devv5*)

- **Process**
  1. Run *load_stage.sql* in the *dev_xyz* schema
  2. QA/QC part 1
  3. Generic update
  4. QA/QC part 2 : semi-automatic process

### Community Contribution Process

- **Prerequisites**
  - Preparation Schemas : *sources*, *dev_xyz*, *devv5*
  - Copies of tables, fully indexed (downloaded from Athena and put into Schema *dev_xyz*, *devv5*)

- **Process**
  1. Run *load_stage.sql* in the *dev_xyz* schema
  2. QA/QC part 1
  3. Generic update
  4. QA/QC part 2 : semi-automatic process

### Community Contribution Process

- **Process**
  1. **Run *load_stage.sql* in the *dev_xyz schema***
     - **If *load_stage* references *devv5* or *sources*, replace them with the names of your schema**

### Community Contribution Process

- **Process**
  1. **Run *load_stage.sql* in the *dev_xyz schema***
     - **If** *load_stage* **references *devv5* or *sources*, replace them with the names of your schema**
     - **Run function** *Vocabulary_pack.SetLatestUpdate.sql*

```
DO $_$
BEGIN
  PERFORM VOCABULARY_PACK.SetLatestUpdate(
  pVocabularyName          => 'vocabulary_id of your vocabulary as in Vocabulary table',
  pVocabularyDate          => 'date of new vocabulary version',
  pVocabularyVersion       => 'name of the vocabulary version, if none use date of the version',
  pVocabularyDevSchema     => 'name of your development schema'
);
END $_$;
```

### Community Contribution Processv

- **Process**
  1. **Run *load_stage.sql* in the *dev_xyz schema***
     - **If *load_stage* references *devv5* or *sources*, replace them with the names of your schema**
     - **Run function** *Vocabulary_pack.SetLatestUpdate.sql*
     - **Run supporting functions to stage tables**

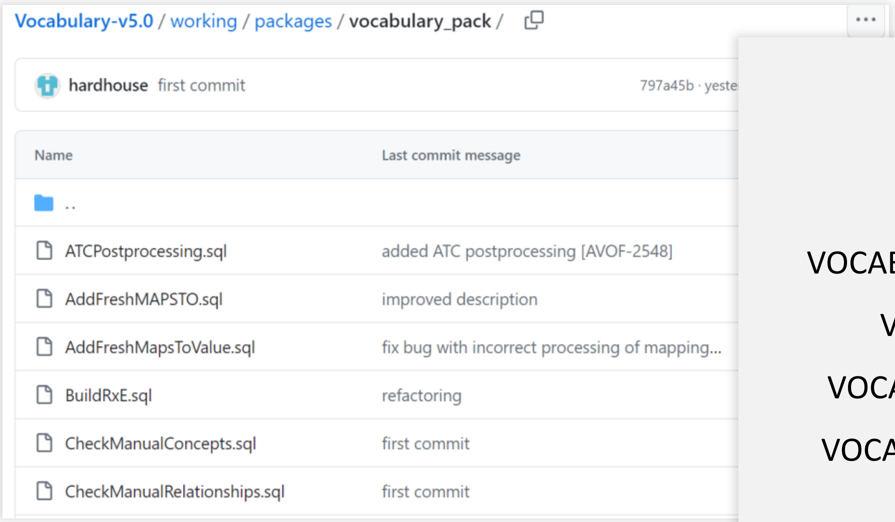


Use these support functions:

VOCABULARY_PACK.ProcessManualRelationships()

VOCABULARY_PACK.AddFreshMAPSTO()

VOCABULARY_PACK.DeprecateWrongMAPSTO()

VOCABULARY_PACK.DeleteAmbiguousMAPSTO()

① You should download function queries you need

③ Run query

④ If you've done it correctly, it should be contained within the *vocabulary_pack* schema functions

### Community Contribution Process

- **Process**
  2. **QA/QC part 1**
     - **As a result of previous step, you will populate stage tables (*concept_stage*, *concept_relationship_stage*, *concept_synonym_stage*, etc)**
     - **Run *qa_tests.check_stage_tables ()* in *create_qa_tests.sql***

```
CREATE OR REPLACE FUNCTION qa_tests.check_stage_tables ()
RETURNS TABLE (
          error_text TEXT,
          rows_count BIGINT
) AS
$BODY$
BEGIN
          RETURN QUERY
          SELECT reason, COUNT(*) FROM (
                    --concept_relationship_stage
                    SELECT
                              CASE WHEN v1.vocabulary_id IS NOT NULL AND
```

This function performs conformance checks and should return no errors.

- If the concept is valid, check valid_end_date = 12/31/2099
- Filed length does not exceed limits in the standard DDL
- There are no duplicates
- Vocabulary_id exist in *VOCABULARY* table

https://github.com/OHDSI/Vocabulary-v5.0/blob/master/working/packages/QA_TESTS/create_qa_tests.sql#L623

### Community Contribution Process

- **Process**

  3. **Generic update**

     - **This function integrates the content of the _*stage* tables into basic tables and assigns concept_ids.**
     - **If you execute this function, you can inspect *CONCEPT* and *CONCEPT_RELATIONSHIP***

```
CREATE OR REPLACE FUNCTION devv5.GenericUpdate (
)
RETURNS void AS
$BODY$
BEGIN
	--1. Prerequisites:
	--1.1 Check vocabulary table, at least one vocabulary must
	PERFORM FROM vocabulary WHERE latest_update IS NOT NULL LI
	IF NOT FOUND THEN
		RAISE EXCEPTION 'At least one vocabulary must ha
			USING HINT = 'Forgot to execute SetLat
	END IF;

	--1.2 Check stage tables for incorrect rows
	DO $$
```

If you need to modify your scripts or stage tables,
you will need to clean stage tables and revert basic tables.

You can easily erase all changes using *fast_recreate_schema.sql*

### Community Contribution Process

- **Process**
  4. **QA/QC part 2 : semi-automatic process**
     - **Execute *get_checks* to check the compliance of resulting basic tables to the OMOP rules**
     - **Run *manual_checks_after_generic* to review manually**

```
CREATE OR REPLACE FUNCTION qa_tests.get_checks (checkid IN INT DEFAULT NULL)
RETURNS TABLE
(
          check_id int4,
          check_name VARCHAR(1000),
          concept_id_1 int4,
          concept_id_2 int4,
          relationship_id VARCHAR(20),
          valid_start_date DATE,
          valid_end_date DATE,
          invalid_reason VARCHAR(1)
)
AS $BODY$
          --relationships cycle
```

* This query is very heavy
It may take a long time to execute.

https://github.com/OHDSI/Vocabulary-v5.0/blob/master/working/packages/QA_TESTS/create_qa_tests.sql#L211

### Community Contribution Process

1. Send the source file + stage tables to Vocabulary Team
2. Create a pull request in Vocabulary GitHub

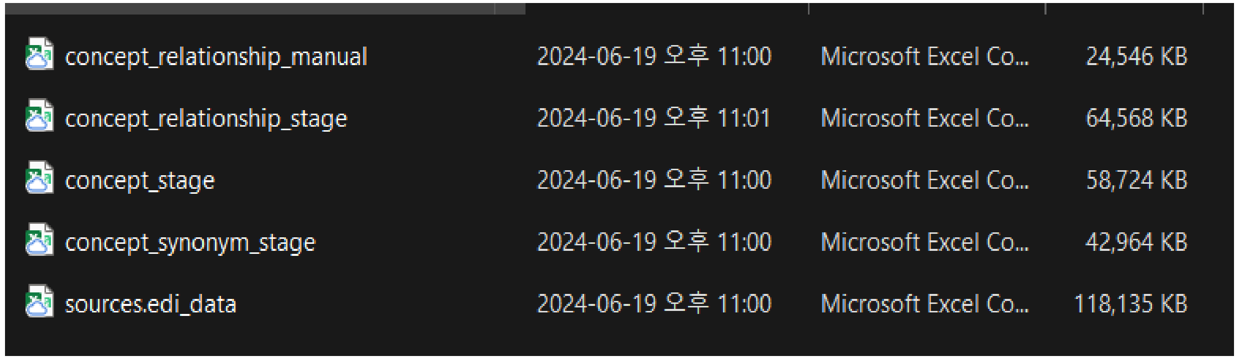


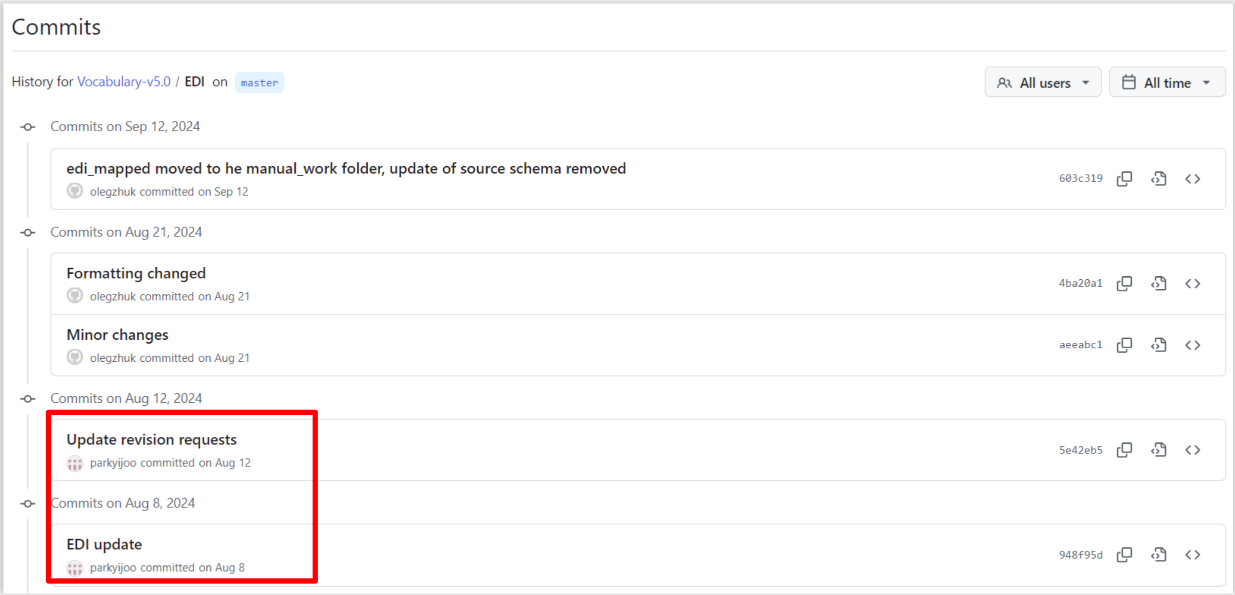

OHDSI
Observational Health Data Sciences and Informatics

### Thank You!
